# Common Barriers to Implementation Across Contexts: Evidence to inform the selection of implementation strategies

**DOI:** 10.1101/2025.09.24.25336521

**Authors:** Luke Wolfenden, Magdalena Wilczynska, Sam McCrabb, Christophe Lecathelinais, Lucy Couper, Tanja Kuchenmuller, Davi Mamblona, Nicole Nathan, Justin Presseau, Natalie Taylor, Rachel Sutherland

## Abstract

**Background:** The implementation of evidence-informed interventions is required to strengthen health systems and improve health outcomes. Identifying implementation barriers underpins evidence-based approaches to do so, but this process is considered complex and time consuming in practice. Understanding whether certain barriers are more common across contexts may guide the development of more effective implementation strategies when comprehensive primary data collection to assess barriers is not feasible.

**Methods:** We conducted a pooled analysis of barrier data from studies that quantitatively assessed implementation barriers using the Theoretical Domains Framework (TDF) survey. To assess barrier frequency aligned to each TDF domain, we calculated the proportion of studies where domain scores were less than four on a standardised 5-point Likert scale. To describe their ‘strength’ we pooled data across studies and reported mean domains scores (lower domain scores represent stronger perceived barriers). Subgroup analyses using pooled domains scores were undertaken to examine differences by population, intervention and geographic characteristics.

**Results:** Data from 42 studies published 2012 to 2024, with a combined sample of 9,809 participants were included in the analysis. In 10 of 14 TDF domains, both mean and median scores were 4 or below, indicating that they were typically perceived as barriers. Four domains had a mean score of 4 or below in >80% of all studies that assessed them - *reinforcement’, ‘environmental context and resources’, social influences’* and *‘behavioural regulation’*. TDF domains with the lowest scores (representing the strongest barriers), were ‘*environmental context and resources*’; ‘*behavioural regulation*’ and ‘*social influences.*’ Few differences (four of 42 statistical comparisons) were found between TDF domain scores and population, intervention and geographic factors.

**Conclusions:** This study identified a set of barriers that appear to be common, and consistent in their perceived strength across a range of population groups, intervention types and geographic localities. The findings provide a basis for those undertaking efforts to improve implementation of evidence-informed health interventions to anticipate types of barriers they may encounter and so, likely strategies that may be needed to address these. This may be beneficial in resource contexts where primary data collection for more comprehensive barrier assessments may not be feasible.

**CONTRIBUTIONS TO THE LITERATURE:**

1. While best practice approaches to the development of effective implementation strategies include assessment of local implementation barriers, many improvement initiatives are undertaken by health organisations or practitioners without the collection of primary data using recommended and valid barrier assessment methods.
2. Systematic reviews suggest similar barriers to implementation of health interventions may exist across a range of contexts. We sought to formally investigate such patterning of barriers, and found evidence of a set of barriers that were both prevalent and salient across contexts.
3. As the tacit knowledge, experience and intuition of health professional are typically the basis of improvement initiatives the study provides some guidance to better help those responsible for improving implementation to anticipate common barriers and devise strategies to address them.

## BACKGROUND

Health innovations capable of reducing much of the burden on patients, communities and health systems currently exist.^1^ Implementation science has emerged as a scientific discipline to facilitate the more rapid and comprehensive implementation of evidence informed interventions and policies into routine health care.^2^ Despite considerable progress in the ‘science’ of implementation, significant gaps between what evidence suggests is effective and what occurs in practice persist.^3^ Approximately 40% of healthcare does not align with contemporary standards of evidence-based practice and may be of low value, or even harmful.^4^ As such, there is an urgent need for more effective approaches to strengthen health systems to support the consistent delivery of evidence-informed high-quality care.

A critical step in the design of effective strategies to improve implementation is assessing and understanding barriers to implementation.^5^ A range of frameworks, methods and tools have been developed to assist in identifying implementation barriers. One of the most commonly used implementation determinants frameworks in implementation science is the Theoretical Domains Framework (TDF).^6, 7^ The TDF includes up to 14 domains, has a validated quantitative survey to measure these,^8^ and published matrices that link these determinants to strategies that could be used to address them to improve implementation.^5, 9–11^ Strategies to improve the implementation of interventions using the TDF have been found to be effective across a range of clinical and community settings and for policies and programs targeting improvement across a variety of health conditions.^12^

The use of determinant frameworks, such as the TDF, has been suggested to improve implementation outcomes by enabling more comprehensive assessments of barriers and precise matching of strategies to address barriers specific to context.^6^ For example, the use of the determinant frameworks has been shown to increase the number and variety of implementation barriers identified by healthcare staff involved in improvement efforts and may improve the effectiveness of implementation initiatives.^6, 13, 14^ Despite these benefits, health care professionals often perceive such frameworks as complex, time consuming, and difficult to apply.^15^ Administering and analysing quantitative surveys, undertaking qualitative research or observations to collect data on implementation determinants – as is recommended by best practice approaches to design effective implementation strategies - is often beyond the resources and capabilities of health services and staff. As such, the use of such frameworks and tools to comprehensively assess barriers are rarely undertaken by staff or organisations seeking to improve health services or implement policy.^12, 16^

In the absence of implementation frameworks, like the TDF, health professionals often rely on their experience and intuition to identify and develop strategies to address implementation barriers. In one study, for example, health professionals intuitively derived implementation strategies aligned with those recommended by theoretical frameworks in only 20% of cases.^17^ Perhaps unsurprisingly, most attempts to improve implementation by health professionals often use a narrow set of implementation strategies (e.g., education material or training,^18^) focused on addressing a relatively small number of perceived barriers such as knowledge or skills which tend to yield only small to moderate benefits.^19^ More practical, theoretically informed approaches to assist health professionals identify relevant implementation barriers are needed.

While barriers to the implementation evidence-informed policies and interventions are context specific, some barriers appear to be more frequently reported than others. For example, barriers classified under the domain ‘environmental context and resources’ such as limited time or resources, has consistently emerged as the most common barrier to implementation in areas such as physical activity policies in schools,^20^ dietary guidelines^21^ or physical activity promotion^22^ in centre based childcare services, planning health care quality,^23^ and clinical behaviour change by primary care practitioners.^24^ This may be, in part, due to more frequent assessment of these factors. Nonetheless, identifying barriers that may be common, or consistently salient, across different contexts may provide a useful resource for healthcare staff or organisations seeking to improve implementation. This may be the case for staff or organisation that have little capacity to apply formal, comprehensive, data driven, and theoretically guided methods of barrier identification. For example, it may support more targeted (e.g., confirmatory) or pragmatic barrier assessments by healthcare providers; support triangulation of local assessments with the broader evidence base; and help select more effective implementation strategies.

## METHODS

### Aim and Study Design

The aim of this study was to: i) describe barriers to the implementation of evidence-informed health policies and interventions, ii) assess the frequency and strength of quantitatively assessed barriers to implementation for each of the 14 domains of the TDF; and iii) assess whether barriers differ by population (the professional group implementing the intervention), intervention, (the type of intervention being implemented) and geographic region (the country where implementation is taking place). We used the Preferred Reporting Items for Systematic Reviews and Meta-Analyses (PRISMA) checklist when writing our report.^25^

### Eligibility Criteria

While there are a range of methods and tools to assess implementation barriers, in this study we focussed on quantitative survey-based methods. We included studies that quantitatively assessed barriers to the implementation of health interventions using instruments aligned with the TDF. Eligible studies were those that used previously validated TDF (and/or modified) based survey instruments. Studies were included if they reported on any TDF domain, using either the 12- or 14-domain version. Studies were included if quantitative measures of TDF constructs enabled data to be pooled for quantitative analyses (mean (M), standard deviation (SD)). We included cross-sectional data. For longitudinal study designs, we included data from baseline assessments only. For intervention trials using comparative designs, we used pre-intervention and/or control condition data where available. We excluded studies where M and SD of TDF constructs (or overall scores) were not reported or could not be calculated with the information provided (e.g., interquartile range: IQR). Additionally, studies or partial results of studies were excluded, if they used inconsistent reporting when calculating TDF domains (e.g. combining different statistical formats such as IQR with M and SD, using mixed scales to assess a particular domain, or omitting critical information). We also excluded protocol papers, systematic reviews, scoping reviews and studies that synthesised qualitative data only, focussed on the implementation of non-health or wellbeing interventions (e.g., manufacturing) or were not published in English.

### Information Sources and Search Strategy

Forward citation searches of key TDF publications^7, 26, 27^ were undertaken to identify eligible publications in April 2024. We also consulted with international experts for any additional potentially eligible papers. The screening process, including title and abstract screening and full-text review of all citations, was conducted by one reviewer (MW). Any uncertainties were resolved through a consensus process with a second senior reviewer (LW). Covidence software^28^ was used to manage the screening process. A detailed summary of the search and screening process is presented in Figure 1.

**Figure 1.**
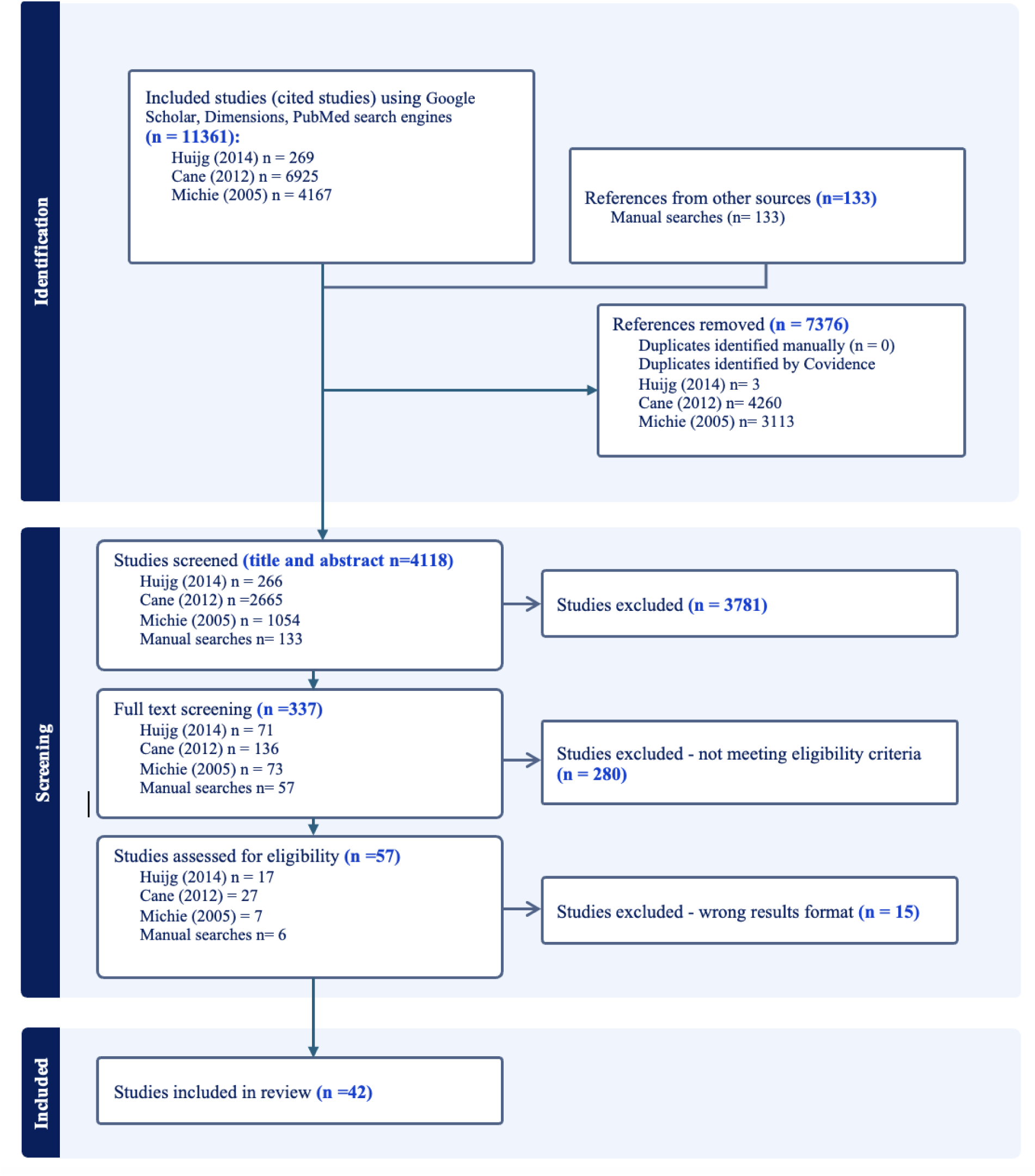
Identification and screening steps taken to identify the eligible quantitative studies.

### Data extraction process

Data were extracted using a pre-defined data extraction form in Microsoft Excel,^29^ developed by the research team. Key study variables which were extracted included:

- Citation details such as publication title, journal, year of publication; and
- Research characteristics and TDF assessments: study aim; design; sample size details about the TDF questionnaire used; Likert scale used; item format; barrier classification method or thresholds; TDF domains assessed; and individual TDF domain scores with measures of variance (e.g., means and standard deviations).

Details of the population or professional group required to implement the intervention (e.g., clinicians, teacher) professions were classified according to the World Health Organization (WHO) ‘Classifying Health Workers’ categorisations.^30^ The location of each study, including the setting, country, region, and corresponding country income classification, was categorised based on the Human Behaviour Change Project’s setting ontology^31^ and the WHO global regions^32^ and income classifications.^33^ Due to the limited variability across the studies (skewed trend) some predefined categories contained insufficient data (e.g., most studies conducted in high-income countries and from the medical/rehabilitation settings). As a result, certain classifications were combined or reduced to allow for meaningful analysis and interpretation.

We classified interventions using the Taxonomy of Health Interventions (informed by Cochrane^34^ and the WHO Best Buys^35^) based on whether they focused on behavioural, psychological or educational based interventions; or those that were medical interventions focused on surgery, physical therapies and pharmacological agents and vaccines.

### Data management and analysis

All analyses were conducted using the statistical program SAS 9.4 and R4.5.1. For studies using terminology that differed from standard TDF domain labels but where the items clearly aligned with the intended domain construct, we applied standardised domain names following the guidelines established by Cane et al. (2012).^7^ To maintain consistency across studies, data using the older 12-domain TDF version^27^ were mapped to the updated 14-domain version.

We calculated M and SD using information provided in the published reports for the overall domain scores. Additionally, where overall scores were not reported but sufficient information was available (i.e. M and SD reported for individual items within TDF domains), we also incorporated these results. In nine studies, where results (e.g., M, SD) were reported for individual items within TDF domains - rather providing a domain score we calculated an overall domain score by averaging the means of items within the domain and deriving a pooled standard deviation using a weighted average. Three studies reported results for multiple professional groups and/or implementation behaviours. In these cases, the data were included as separate entities as data was available for different behaviours or professional groups (Table 1). Studies that included TDF domain scores using a single item were included. We standardised all TDF domain scores to a 5-point Likert scale.

**Table 1.**
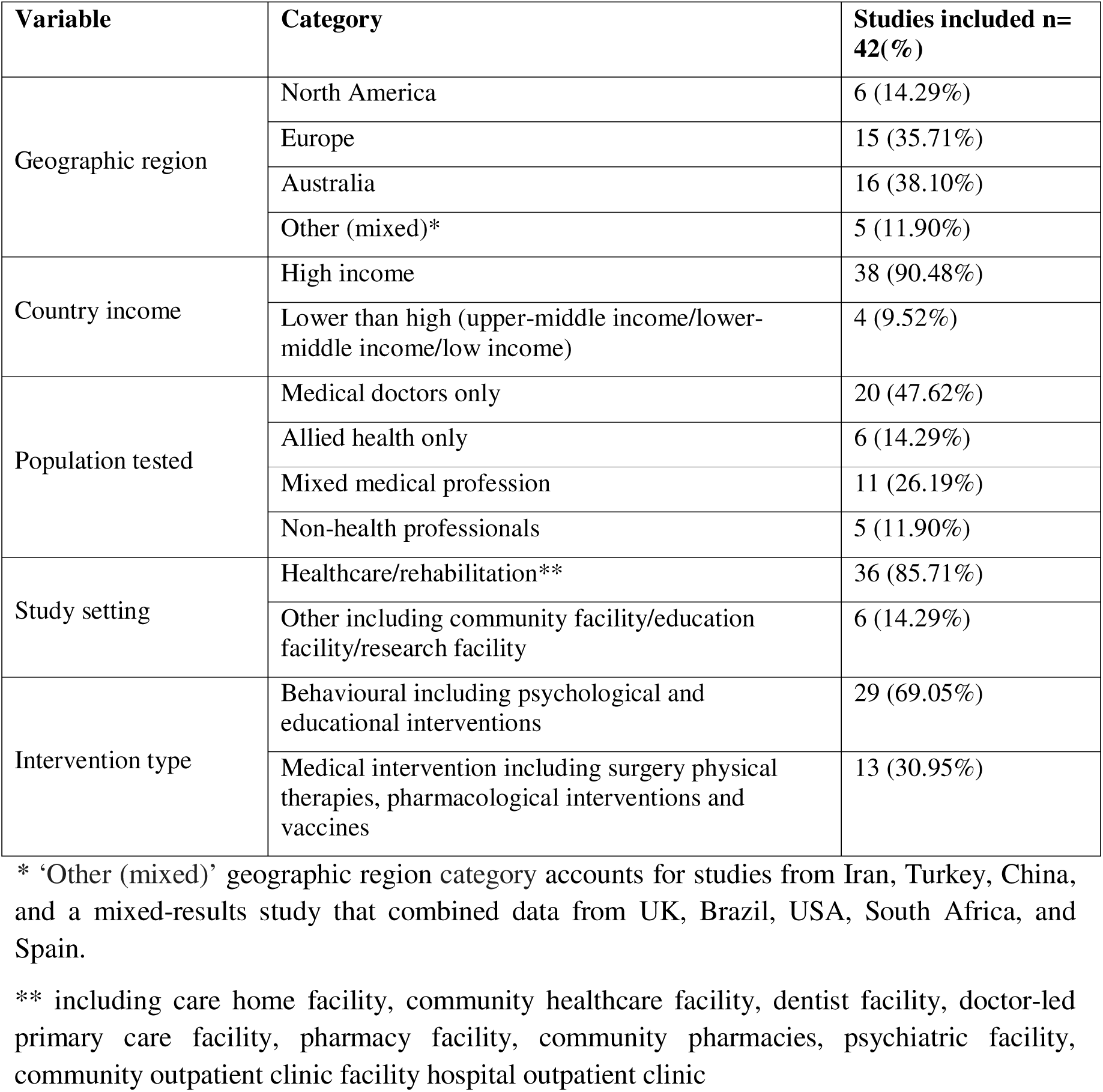
Baseline demographics of the included studies (n=42).

| **Variable** | **Category** | **Studies included n=42(%)** |
| --- | --- | --- |
| Geographic region | North America | 6 (14.29%) |
|  | Europe | 15 (35.71%) |
|  | Australia | 16 (38.10%) |
|  | Other (mixed)* | 5 (11.90%) |
| Country income | High income | 38 (90.48%) |
|  | Lower than high (upper-middle income/lower-middle income/low income) | 4 (9.52%) |
| Population tested | Medical doctors only | 20 (47.62%) |
|  | Allied health only | 6 (14.29%) |
|  | Mixed medical profession | 11 (26.19%) |
|  | Non-health professionals | 5 (11.90%) |
| Study setting | Healthcare/rehabilitation** | 36 (85.71%) |
|  | Other including community facility/education facility/research facility | 6 (14.29%) |
| Intervention type | Behavioural including psychological and educational interventions | 29 (69.05%) |
|  | Medical intervention including surgery physical therapies, pharmacological interventions and vaccines | 13 (30.95%) |

\* 'Other (mixed)' geographic region category accounts for studies from Iran, Turkey, China, and a mixed-results study that combined data from UK, Brazil, USA, South Africa, and Spain.
\*\* including care home facility, community healthcare facility, dentist facility, doctor-led primary care facility, pharmacy facility, community pharmacies, psychiatric facility, community outpatient clinic facility hospital outpatient clinic

Descriptive statistics were used to summarise study characteristics. To assess the frequency of barriers to implementation for each TDF domain, we calculated the proportion of studies where domain scores were four or less on a 5-point Likert scale. A previous study has recommended and applied this cut point to identify ‘barriers’ to implementation.^36^ To ascertain the ‘strength’ of a barrier for each domain, we pooled mean scores across all studies, using a random effects meta-analysis. Lower domain scores represent stronger perceived barriers to implementation. Point estimates of pooled domain scores were then used to rank domains from strongest to weakest perceived barriers. Comparisons of the confidence intervals around barrier point estimates were used to infer statistical differences in domain scores (strength). As authors of included studies may assess only domains considered most likely to represent a barrier; most relevant to their context; or those that more easily addressed (selection bias) we conducted sensitivity analyses including only studies reporting all 14 TDF domains.

To determine whether barriers differ by subgroup, analyses using pooled domains scores were undertaken by population (medical, allied health/mixed, non-health professionals), intervention type (medical/surgery, behavioural/psychological) or geographic region (North America, Europe, Australia, Other [mixed]). Statistical tests for pooled estimate are reported as well as pooled estimates and 95% confidence intervals for subgroups examined. Statistical tests were two tailed with an alpha of 0.05. While we had intended to report findings by WHO regions, and income levels, the concentration of included studies in a relatively small number of countries precluded this. Instead, we present subgroups in more specific geographic groupings that reflect where the research was undertaken.

## RESULTS

### Results Summary

After screening 4,118 titles, a total of 42 studies met the eligibility criteria, with a combined sample of 9,809 participants (Figure 1). Most studies were conducted in Europe (35.7%) and Australia (38.1%) and originated from high-income countries (90.5%). Less than half of the studies focused on medical doctors (47.6%) and were typically set in healthcare or rehabilitation contexts (85.7%). Eight of the 42 studies report data across 14 TDF domains. The primary focus of implementation efforts was behavioural, psychological, or educational interventions (69%). Further baseline characteristics are detailed in Table 1.

Most of the studies (55%) were published in the past four years, publications ranged from 2012–2024, with 2021 accounting for the highest number of publications included in this review (21%). The sample size ranged from 8 – 1,090. Only 19% of studies (n= 8) assessed implementation barriers across all 14 TDF domains. Domains most frequently examined included *‘beliefs about capabilities’, ‘beliefs about consequences’, ‘knowledge’*, *‘skills’*, *’social/professional role and identity’*, *‘environmental context and resources’*, and *‘social influences’*. In contrast, *‘optimism’* and *‘reinforcement’* were least commonly assessed (Table 2).

**Table 2.** TDF domain scores across all studies included (n=42) and number (%) of studies where domain scores were = 4 or less.

| **TDF Domain** | **N<sup>a</sup>** | **Mean (SD)<sup>b</sup>** | **Median** | **IQR** | **Min; Max** | **Number (%) of studies with mean <4 (barriers)** |
| --- | --- | --- | --- | --- | --- | --- |
| 1. Knowledge | 36 | 4.03 (0.50) | 4.04 | 3.64 - 4.45 | 3.12; 4.87 | 16 (44.44%) |
| 2. Skills<sup>*</sup> | 35 | 3.79 (0.60) | 3.85 | 3.36 - 4.28 | 2.39; 4.76 | 21 (60.00%) |
| 3. Social/professional role and identity | 35 | 4.16 (0.40) | 4.17 | 3.91 - 4.53 | 3.16; 4.79 | 12 (34.29%) |
| 4. Beliefs about capabilities<sup>*</sup> | 38 | 3.76 (0.51) | 3.73 | 3.30 - 4.12 | 2.59; 4.74 | 25 (65.79%) |
| 5. Optimism<sup>*</sup> | 14 | 4.00 (0.42) | 3.95 | 3.78 - 4.46 | 3.23; 4.53 | 9 (64.29%) |
| 6. Beliefs about consequences | 36 | 4.05 (0.50) | 4.17 | 3.72 - 4.35 | 2.59; 4.92 | 15 (41.67%) |
| 7. Reinforcement<sup>*</sup> | 13 | 3.60 (0.46) | 3.75 | 3.48 - 3.82 | 2.33; 4.20 | 12 (92.31%) |
| 8. Intentions | 27 | 4.03 (0.42) | 4.07 | 3.78 - 4.33 | 2.89; 4.70 | 10 (37.04%) |
| 9. Goals<sup>*</sup> | 29 | 3.66 (0.73) | 3.63 | 3.38 - 4.25 | 1.80; 4.84 | 20 (68.97%) |
| 10. Memory, attention and decision processes<sup>*</sup> | 26 | 3.64 (0.47) | 3.64 | 3.45 - 4.01 | 2.44; 4.47 | 19 (73.08%) |
| 11. Environmental context and resources<sup>*</sup> | 36 | 3.40 (0.59) | 3.34 | 3.13 - 3.84 | 1.87; 4.43 | 29 (80.56%) |
| 12. Social influences<sup>*</sup> | 36 | 3.55 (0.50) | 3.46 | 3.19 - 3.80 | 2.20; 4.62 | 30 (83.33%) |
| 13. Emotion<sup>*</sup> | 29 | 3.65 (0.63) | 3.70 | 3.37 - 4.07 | 2.13; 4.74 | 20 (68.97%) |
| 14. Behavioural regulation<sup>*</sup> | 24 | 3.54 (0.51) | 3.58 | 3.17 - 3.91 | 2.69; 4.38 | 20 (83.33%) |

a Number of included studies included in this review that assessed the specific TDF domain.
\* Mean score of 4 or less indicates that domain was identified as a barrier.

### Frequency and strength of barriers

Across the 14 TDF domains, ten had both mean and median scores below 4 (Table 2), indicating that they were typically (on average) perceived as barriers to implementation. These domains included: ‘*skills,’ ‘beliefs about capabilities,’ ‘reinforcement,’ ‘goals,’ ‘memory, attention and decision processes,’ ‘environmental context and resources,’ ‘optimism,’ ‘social influences,’ ‘emotion,’* and *‘behavioural regulation.’* In sensitivity analyses using data only from eight studies reporting scores for all 14 domains*, ‘knowledge,’* but not *‘skills’* was identified as a barrier based on mean and median scores (Appendix 1). No other differences in barriers classifications were identified in sensitivity analysis.

Ten of the TDF domains had barriers mapped to them in more than half of the studies that assessed them. Barriers were most frequently mapped to four domains (*‘reinforcement,’ ‘environmental context and resources,’ social influences,’* and *‘behavioural regulation’*) each reported in more than 80% of the relevant studies. An additional domain, *‘memory, attention and decision processes,’* was captured barriers in 73% of studies. In sensitivity analyses, these five domains were also identified as a barrier in 75% or more of the included studies (Appendix 1). The domain most frequently associated with barriers was *‘reinforcement,’* with over 92% of studies reporting barriers with this domain. In contrast, barriers related to *‘social/professional role and identity’* were least frequent, reported in only 34% of the studies. These findings were also consistent with that of sensitivity analysis (Appendix 1).

Based on meta-analysis the, TDF domains with the lowest pooled domain scores, suggesting they represent the strongest barriers to implementation were (in rank order) *‘environmental context and resources,’ ’behavioural regulation,’ ‘social influences,’ ‘reinforcement,’* and *‘memory, attention and decision processes’* (Table 3). In sensitivity analyses, the same five domains emerged (in rank order) *‘environmental context and resources,’ ‘memory, attention and decision processes,’ ‘reinforcement,’ ‘social influences,’ and ’behavioural regulation’* (Appendix 1).

**Table 3.** The meta-analysis results of TDF domains summarised as the frequency, and pooled mean results per each TDF domain tested.

| **TDF domains** | **Number of included studies** | **Total sample tested** | **Meta-analysis Estimate CI (95%)** | **Rank (1= domain representing the strongest barrier)** | **Non overlapping confidence intervals between TDF domains scores** |
| --- | --- | --- | --- | --- | --- |
| 1. Knowledge | 36 | 9216 | 4.03 [3.86; 4.19] | 12 | TDF Domain 1 → Domain 7, 10, 11, 12, 14 |
| 2. Skills* | 35 | 8878 | 3.79 [3.59; 3.99] | 9 | TDF Domain 2 → Domain 3 |
| 3. Social/professional role and identity | 35 | 8852 | 4.16 [4.03; 4.29] | 14 | TDF Domain 3 → Domain 2, 4, 7, 9, 10, 11, 12, 13, 14 |
| 4. Beliefs about capabilities* | 38 | 7225 | 3.77 [3.61; 3.93] | 8 | TDF Domain 4 → Domain 3, 11 |
| 5. Optimism* | 14 | 2690 | 4.00 [3.78; 4.22] | 10 | TDF Domain 5 → Domain 11, 12, 14 |
| 6. Beliefs about consequences | 36 | 9001 | 4.05 [3.89; 4.21] | 13 | TDF Domain 6 → Domain 7, 10, 11, 12, 13, 14 |
| 7. Reinforcement* | 13 | 1907 | 3.60 [3.35; 3.84] | 4 | TDF Domain 7 → Domain 1, 3, 6, 8, |
| 8. Intentions | 27 | 5817 | 4.02 [3.86; 4.19] | 11 | TDF Domain 8 → Domain 7, 10, 11, 12, 14 |
| 9. Goals* | 29 | 5961 | 3.66 [3.40; 3.93] | 7 | TDF Domain 9 → Domain 3 |
| 10. Memory, attention and decision processes<sup>a*</sup> | 26 | 5523 | 3.65 [3.47; 3.83] | 5 | TDF Domain 10 → Domain 1, 3, 6, 8, |
| 11. Environmental context and resources* | 36 | 9051 | 3.40 [3.21; 3.60] | 1 | TDF Domain 11 → Domain 1, 3, 4, 5, 6, 8 |
| 12. Social influences* | 36 | 9189 | 3.55 [3.39; 3.71] | 3 | TDF Domain 12 → Domain 1, 3, 5, 6, 8 |
| 13. Emotion<sup>a*</sup> | 29 | 8831 | 3.65 [3.42; 3.88] | 6 | TDF Domain → Domain 3, 6 |
| 14. Behavioural regulation* | 24 | 4914 | 3.54 [3.33; 3.74] | 2 | TDF Domain → Domain 1, 3, 5, 6, 8 |

\* indicates a domain with a pooled mean score of 4 or less, indicating a barrier. <sup>a</sup> Given the tighter confidence interval around 'Memory attention and decision processes' when compared to 'Emotion' which has the same estimate, Memory, attention and decision processes' was ranked 5<sup>th</sup> and Emotion 6<sup>th</sup>.

**Table 4.**
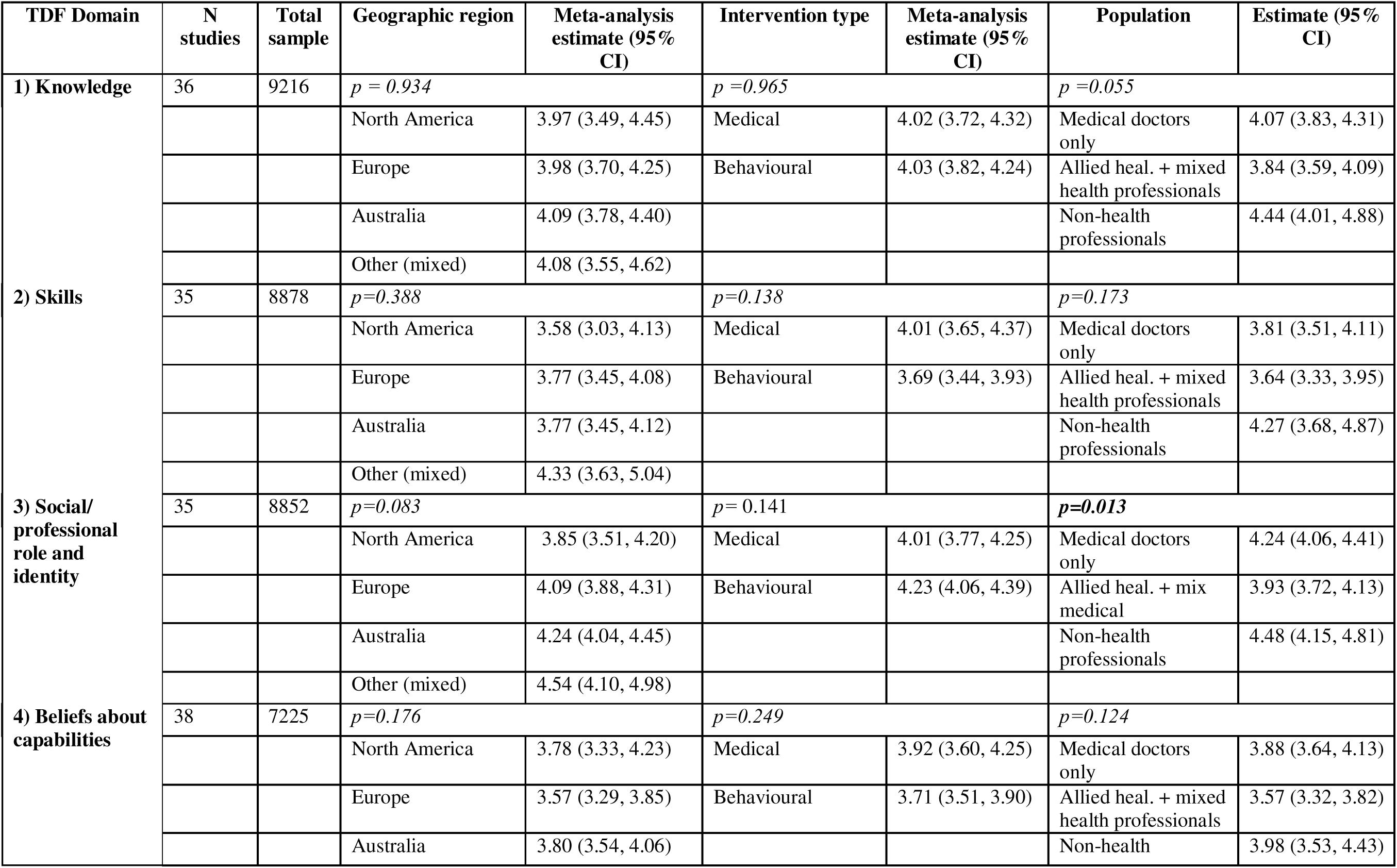
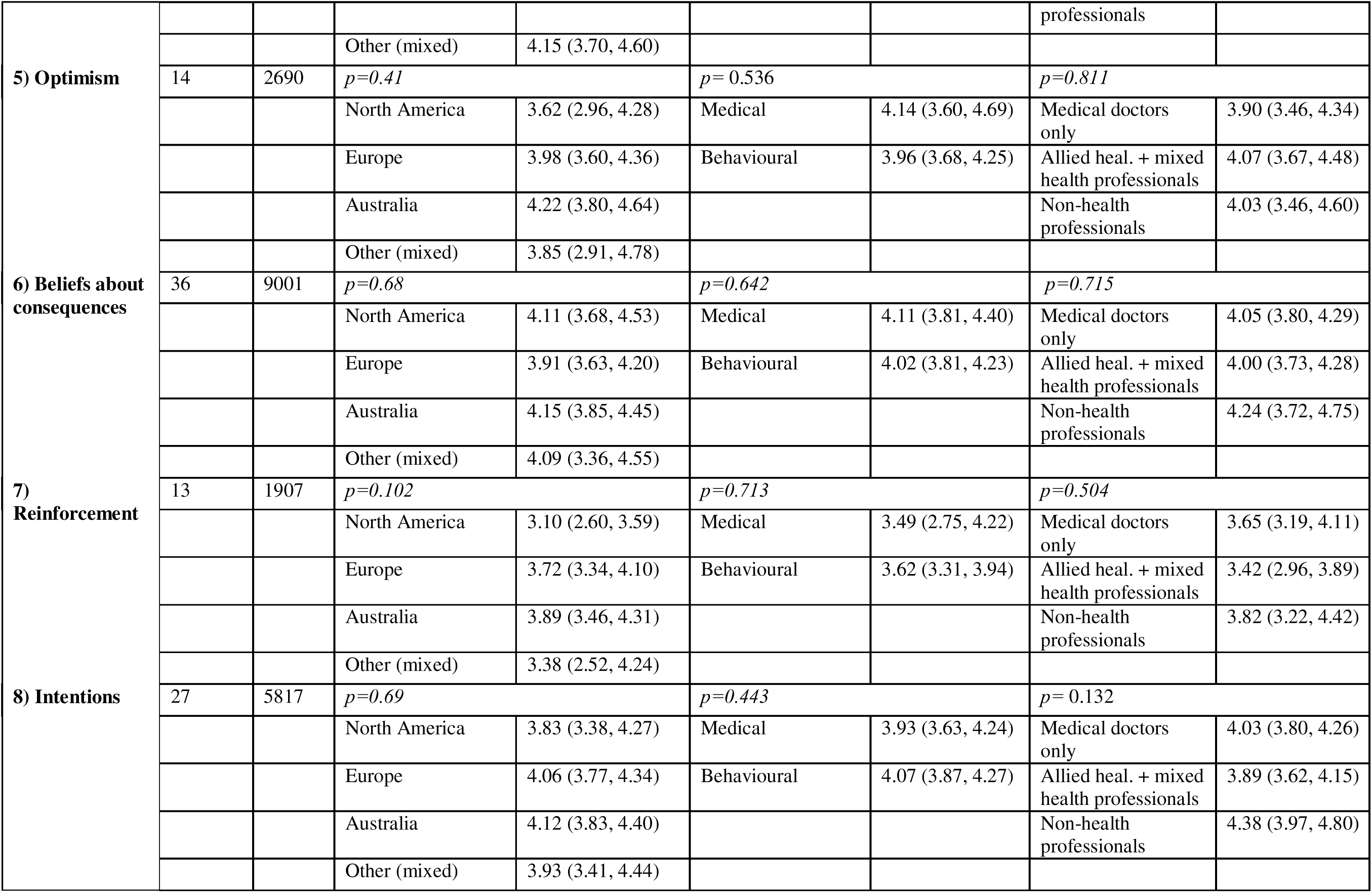
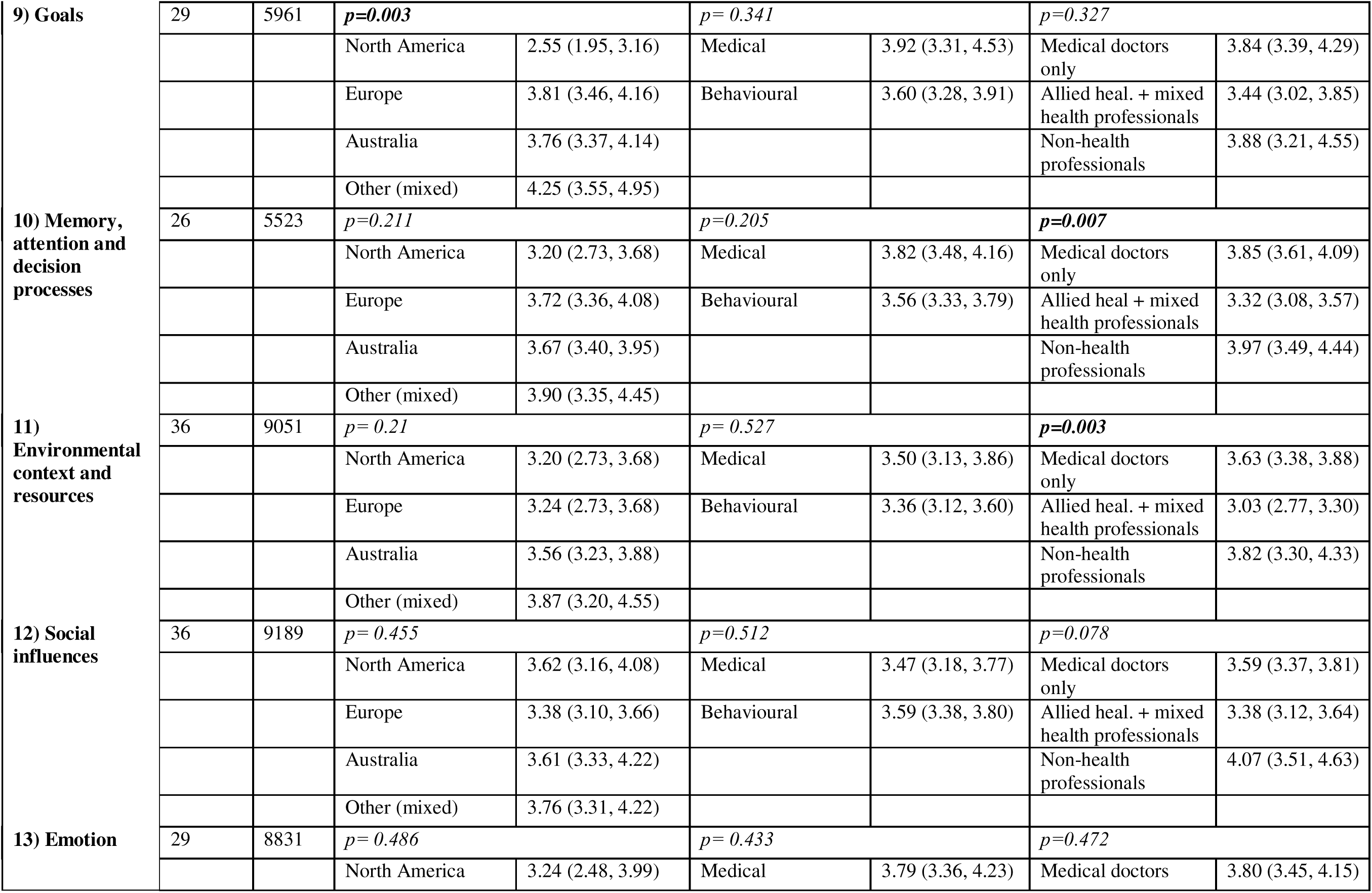
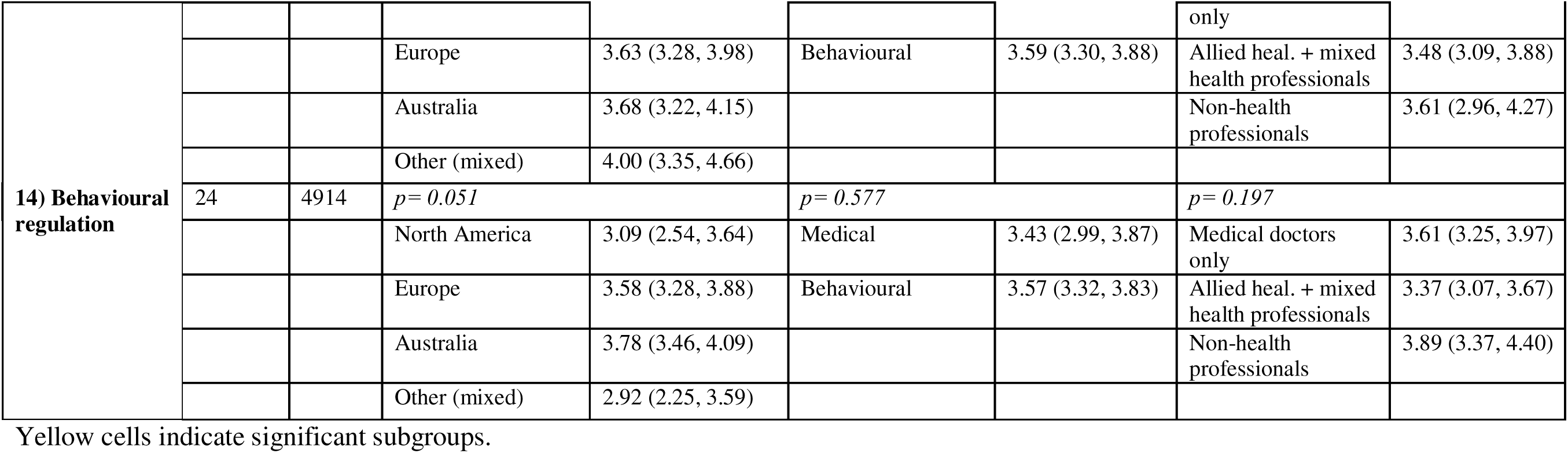
Subgroup analyses of TDF domain scores.

The confidence intervals around pooled estimates of TDF scores did not overlap for a several domains, indicating statistically significant differences (p<0.05). For example, pooled means for domains were significantly different between the top three (‘*environmental context and resources,’ ’behavioural regulation,’ and social influences’*) and bottom four (*‘social/professional role and identify,’ ‘beliefs about consequences,’ ‘knowledge,’ and ‘intentions’*) ranked domains.

Estimates from meta-analysis of domains scores using all available data were similar to those undertaken as part of sensitivity analysis (Appendix 1) (median: 0.06; mean difference range: 0.01-0.27).

### Subgroup analyses by population, intervention and geographic factors

We conducted 42 statistical comparisons for interaction between TDF domain scores and population, intervention type and geographic region. In only four instances were these statistically significant. For the population, significant interactions were found for the TDF domains of ‘*social/professional role and identity,’ ‘memory, attention and decision processes’* and *‘environmental context and resources.’* In each case, mean subgroup scores were lowest among allied health/mixed health profession subgroup, compared with medical doctors and non-health professional subgroups. For geographic region, significant interactions were found for two domains, ‘*goals’* where scores were lowest in studies undertaken in North America compared with those in Europe, Australia or other countries. No significant interactions between intervention type subgroups (medical interventions vs behavioural interventions) across any of 14 TDF domains.

## DISCUSSION

These findings contribute to the broader field of evidence-informed health policy and practice by offering actionable insights into implementation barriers. This study sought to assess the frequency and perceived strength of barriers to implementation of health interventions, and to examine whether barriers differed by population, intervention, or geographic factors. Using data from 9,809 participants across 42 studies, we identified consistent patterns in the types of barriers based on theoretically aligned measures of implementation determinants. While most (10) of the TDF domains had barriers that were mapped to them in more than half of the studies that assessed them, four were consistently reported in almost all (>80%). Furthermore, while differences in strength (or importance) of barriers were identified overall, we found that barriers rarely differed substantively by the population, intervention or geographic characteristics assessed.

Barriers were most frequently mapped to the domains of *‘Reinforcement,’* ‘*environmental context and resources,’ ‘social influences,’* and *‘Behavioural regulation.’* These four domains were also those that were reported as having the lowest domains scores, and so, thought to be the most salient (strongest) impediments to implementation. Some, including *‘social influences,’* are also barriers that healthcare professionals find challenging to identify theory aligned strategies to address.^37^ The findings suggest those responsible for implementing health interventions could anticipate the need to address such barriers and could benefit from clear specification of evidence-informed strategies that could do so. For example, strategies that may address barriers associated with *‘social influences’* could consider the use of peer modelling, opinion leaders, or leveraging social norms. Similarly, strategies to address barriers pertaining to ‘*behavioural regulation’* could involve supporting health care professionals with goal setting or action planning. The study findings also suggest that research to optimise the effectiveness of strategies to address such common, salient barriers could be particularly beneficial.

Interestingly, we found barriers related to the domains of *‘knowledge’* and *‘skills,’* were less commonly identified (44-60% of studies) and were ranked lower in terms of their strength or salience (=11^th^ and 8^th^ respectively). Systematic reviews have identified that strategies to build knowledge and skills such as training, educational meetings or outreach visits are among the most frequently used strategies to improve implementation.^38, 39^ They are also those most likely suggested by practitioners to improve implementation. The findings suggests that the use of such strategies to improve implementation are, in many instances, insufficient when used in isolation.^19, 38, 40^

We found few differences in TDF domains scores when comparing studies by intervention type, population, or geographic locality of the study. For example, no significant (subgroup) differences in domains scores between medically focused interventions and behavioural/psychological focused interventions. This may reflect the relatively broad categories used in subgroup analyses; that quantitative measures used do not capture contextual complexity, such as the potential overlapping nature of barriers; or other biases.^17^ Nonetheless the consistency in findings across diverse contexts provide a basis for those undertaking efforts to improve the implementation of health intervention to anticipate the types of barriers, and the likely strategies that will be required to address them. Specifically, the findings suggest that regardless of the factors examined in this study that certain types of barriers, specifically those related to the domains of ‘*behavioural regulation,’ ‘social influences,’ ‘reinforcement,’ ‘social/professional role and identity,’* are common across a range of implementation contexts. Understanding this may help those responsible for implementation to anticipate and plan for such barriers, and may be particularly valuable if more comprehensive, formal barriers assessment are not feasible. Resources and practical examples of strategies that have been successfully applied to address these barriers may be particularly beneficial for busy and resource limited health services and staff to support their improvement efforts.

There are a number of limitations of the study that require consideration when interpretating the findings. First, the study used data collected from items assessing constructs of a single theoretical framework – the TDF. While the TDF is a widely used framework in implementation science, other frameworks, such as the Consolidated Framework for Implementation Research (CFIR) includes more specific, refined and multi-level determinants.^41^ CFIR theorised determinants for implementation may differ in terms of their frequency, strength and extent across contextual factors assessed in this study. Second, while the study draws on data from almost 10,000 participants, it was extracted from a relatively small number of studies (n=42) that provided limited contextual variability. For example, most studies were undertaken in a small number of countries. This did not allow assessment of the extent to which barriers may vary between high-, low- and middle-income countries. Third, analyses may have also be at risk of confounding. For example, assessment of barriers may be more likely in studies where these barriers were hypothesised to exist. Sensitivity analyses including only studies including assessment of all 14 domains, however, reported similar studies to those undertaken using all available data from all included studies. Nonetheless further research is required to verify these findings, particularly among domains such as *‘reinforcement’* that were less frequently examined by the included studies but frequently identified as barriers in those that did. Fourth, while cross-sectional surveys provide useful insights to identify perceived barriers, prospective associations of these with measures of implementation is required to verify them as determinants. Cross sectional, quantitative surveys also provide limited depth in understanding the complexity of implementation barriers that qualitative or mixed method approaches are able to elicit.

## CONCLUSIONS

Notwithstanding these limitations, the study provides valuable information to guide future implementation research in practice. The study describes patterns in implementation barriers that may help those responsible for improving professional practice anticipate factors that may impede these efforts. In doing so, it may aid the planning and development of effective implementation strategies. The findings do not negate the importance of more rigorous approaches to barrier identification, particularly localised assessment of implementation barriers through quantitative, qualitative or other means; or the tacit knowledge and expertise of health professionals. They may, however, provide an important adjunct to such data collection, or useful input where more comprehensive efforts are not feasible.

## LIST OF ABBREVIATIONS

Term: Abbreviation
Theoretical Domains Framework: TDF
Mean: M
Standard Deviation: SD
Interquartile Range: IQR
World Health Organization: WHO
Consolidated Framework for Implementation Research: CFIR

## DECLARATIONS

### Ethics approval and consent to participate

Not applicable

### Consent for publication

Not Applicable

### Availability of data and materials

The datasets used and/or analysed during the current study are available from the corresponding author on reasonable request.

### Authors’ contributions

LW conceptualised and managed the study. MW lead the identification of included studies and data extraction. CL conducted analysis along with MW and LW led the drafting the manuscript supported by SMc and LC. MW, SMc, CL, LC, TK, DM, NN, JP, NT and RS all read, provided critical comments and approved the final article for submission.

## Data Availability

All data produced in the present study are available upon reasonable request to the authors

## Acknowledgements

Not applicable

