## Appendix 1 for "Common Barriers to Implementation Across Contexts: Evidence to inform the selection of implementation strategies"

**Supplementary Table 1.** The results summary of TDF domains across all studies included (n=42) presented as Means, SDs, IQR, Minimum and Max values and number (%) of studies with mean smaller than 4 (classified as barriers).

|  | **New Results** | | | | | **Original Results** | | | | |
| --- | --- | --- | --- | --- | --- | --- | --- | --- | --- | --- |
| **TDF Domain** | **Mean (SD)^b^** | **Median** | **IQR** | **Min; Max** | **Number (%) of studies with mean <4 (barriers)** | **Mean (SD)^b^** | **Median** | **IQR** | **Min; Max** | **Number (%) of studies with mean <4 (barriers)** |
| 1. Knowledge | 3.90 (0.62) | 3.94 | 3.34 - 4.48 | 3.12; 4.61 | 4 (50.00%) | 4.03 (0.50) | 4.04 | 3.64 - 4.45 | 3.12; 4.87 | 16 (44.44%) |
| 2. Skills | 4.04 (0.40) | 4.03 | 3.79 - 4.43 | 3.36; 4.47 | 4 (50.00%) | 3.79 (0.60) | 3.85 | 3.36 - 4.28 | 2.39; 4.76 | 21 (60.00%) |
| 3. Social/professional role and identity | 4.20 (0.37) | 4.10 | 4.02 - 4.48 | 3.63; 4.78 | 1 (12.50%) | 4.16 (0.40) | 4.17 | 3.91 - 4.53 | 3.16; 4.79 | 12 (34.29%) |
| 4. Beliefs about capabilities | 3.82 (0.48) | 3.73 | 3.46 - 4.26 | 3.17; 4.48 | 5 (62.50%) | 3.76 (0.51) | 3.73 | 3.30 - 4.12 | 2.59; 4.74 | 25 (65.79%) |
| 5. Optimism | 3.99 (0.46) | 3.89 | 3.73 - 4.46 | 3.23; 4.50 | 5 (62.50%) | 4.00 (0.42) | 3.95 | 3.78 - 4.46 | 3.23; 4.53 | 9 (64.29%) |
| 6. Beliefs about consequences | 4.32 (0.24) | 4.34 | 4.16 - 4.47 | 3.93; 4.67 | 1 (12.50%) | 4.05 (0.50) | 4.17 | 3.72 - 4.35 | 2.59; 4.92 | 15 (41.67%) |
| 7. Reinforcement | 3.61 (0.57) | 3.81 | 3.49 - 3.88 | 2.33; 4.20 | 7 (87.50%) | 3.60 (0.46) | 3.75 | 3.48 - 3.82 | 2.33; 4.20 | 12 (92.31%) |
| 8. Intentions | 4.14 (0.49) | 4.19 | 3.86 - 4.54 | 3.25; 4.70 | 2 (25.00%) | 4.03 (0.42) | 4.07 | 3.78 - 4.33 | 2.89; 4.70 | 10 (37.04%) |
| 9. Goals | 3.69 (0.58) | 3.66 | 3.24 - 4.12 | 2.90; 4.55 | 6 (75.00%) | 3.66 (0.73) | 3.63 | 3.38 - 4.25 | 1.80; 4.84 | 20 (68.97%) |
| 10. Memory, attention and decision processes | 3.55 (0.61) | **3.59** | 3.26 - 3.87 | 2.44; 4.47 | 6 (75.00%) | 3.64 (0.47) | 3.64 | 3.45 - 4.01 | 2.44; 4.47 | 19 (73.08%) |
| 11. Environmental context and resources | 3.37 (0.54) | **3.24** | 3.18 - 3.60 | 2.53; 4.43 | 7 (87.50%) | 3.40 (0.59) | 3.34 | 3.13 - 3.84 | 1.87; 4.43 | 29 (80.56%) |
| 12. Social influences | 3.61 (0.68) | 3.70 | 3.49 - 3.83 | 2.20; 4.61 | 7 (87.50%) | 3.55 (0.50) | 3.46 | 3.19 - 3.80 | 2.20; 4.62 | 30 (83.33%) |
| 13. Emotion | 3.73 (0.53) | 3.83 | 3.41 - 4.12 | 2.73; 4.41 | 5 (62.50%) | 3.65 (0.63) | 3.70 | 3.37 - 4.07 | 2.13; 4.74 | 20 (68.97%) |
| 14. Behavioural regulation | 3.66 (0.52) | **3.60** | 3.36 - 4.10 | 2.83; 4.33 | 6 (75.00%) | 3.54 (0.51) | 3.58 | 3.17 - 3.91 | 2.69; 4.38 | 20 (83.33%) |

TDF domains classified as barriers with a mean weighted average below 4 and highlighted in orange.

a Number of included studies included in this review that tested the specific TDF domain.

b Mean score >4 indicates that domain was identified as a barrier

**Supplementary table 2.** The meta-analysis results of TDF domains summarised as the frequency and pooled mean results per each TDF domain tested

|  | **Subgroup^b^** | | | **Whole sample^b^** | | |
| --- | --- | --- | --- | --- | --- | --- |
| **TDF domains** | **Meta-analysis Estimate CI (95%)** | **Rank (1= domain representing the strongest barrier)** | **Non overlapping confidence intervals between TDF domains scores** | **Meta-analysis Estimate CI (95%)** | **Rank (1= domain representing the strongest barrier)** | **Non overlapping confidence intervals between TDF domains scores** |
| 1. Knowledge | 3.91 [3.48; 4.33] | 9 |  | 4.03 [3.86; 4.19] | 12 | TDF Domain 1 🡪 Domain 7, 10, 11, 12, 14 |
| 2. Skills | 4.04 [3.77; 4.32] | 11 | TDF Domain 2 🡪 Domain 11 | 3.79 [3.59; 3.99] | 9 | TDF Domain 2 🡪 Domain 3 |
| 3. Social/professional role and identity | 4.20 [3.94; 4.47] | 13 | TDF Domain 3 🡪 Domain 11 | 4.16 [4.03; 4.29] | 14 | TDF Domain 3 🡪 Domain 2, 4, 7, 9, 10, 11, 12, 13, 14 |
| 4. Beliefs about capabilities | 3.81 [3.48; 4.15] | 8 | TDF Domain 4 🡪 Domain 6 | 3.77 [3.61; 3.93] | 8 | TDF Domain 4 🡪 Domain 3, 11 |
| 5. Optimism | 3.99 [3.67; 4.30] | 10 |  | 4.00 [3.78; 4.22] | 10 | TDF Domain 5 🡪 Domain 11, 12, 14 |
| 6. Beliefs about consequences | 4.32 [4.16; 4.89] | 14 | TDF Domain 6 🡪 Domain 4, 7, 9, 10, 11, 12, 13, 14 | 4.05 [3.89; 4.21] | 13 | TDF Domain 6 🡪 Domain 7, 10, 11, 12, 13, 14 |
| 7. Reinforcement | **3.61 [3.22; 4.01]** | 3 | TDF Domain 7 🡪 Domain 6 | **3.60 [3.35; 3.84]** | 4 | TDF Domain 7 🡪 Domain 1, 3, 6, 8, |
| 8. Intentions | 4.14 [3.80; 4.48] | 12 | TDF Domain 8 🡪 Domain 11 | 4.02 [3.86; 4.19] | 11 | TDF Domain 8 🡪 Domain 7, 10, 11, 12, 14 |
| 9. Goals | 3.69 [3.29; 4.09] | 6 | TDF Domain 9 🡪 Domain 6 | 3.66 [3.40; 3.93] | 7 | TDF Domain 9 🡪 Domain 3 |
| 10. Memory, attention and decision processes^a^ | **3.55 [3.12; 3.97]** | 2 | TDF Domain 10 🡪 Domain 6 | 3.65 [3.47; 3.83] | 5 | TDF Domain 10 🡪 Domain 1, 3, 6, 8, |
| 11. Environmental context and resources | **3.38 [3.00; 3.76]** | 1 | TDF Domain 11 🡪 Domain 2, 3, 6, 8 | **3.40 [3.21; 3.60]** | 1 | TDF Domain 11 🡪 Domain 1, 3, 4, 5, 6, 8 |
| 12. Social influences | **3.62 [3.16; 4.08]** | 4 | TDF Domain 12 🡪 Domain 6 | **3.55 [3.39; 3.71]** | 3 | TDF Domain 12 🡪 Domain 1, 3, 5, 6, 8 |
| 13. Emotion^a^ | 3.73 [3.35; 4.10] | 7 | TDF Domain 🡪 Domain 6 | 3.65 [3.42; 3.88] | 6 | TDF Domain 🡪 Domain 3, 6 |
| 14. Behavioural regulation | **3.65 [3.29; 4.01]** | 5 | TDF Domain 🡪 Domain 6 | **3.54 [3.33; 3.74]** | 2 | TDF Domain 🡪 Domain 1, 3, 5, 6, 8 |

TDF domains classified as barriers with a mean weighted average below 4 and highlighted in orange.

^a^ Given the tighter confidence interval around ‘Memory attention and decision processes’ when compared to ‘Emotion’ which has the same estimate, Memory, attention and decision processes’ was ranked 5^th^ and Emotion 6^th^.

^b^ The subgroup includes 8 studies, and a population of 1,207. The whole sample includes 42 studies and a population of 9,8009.
